# Condition-Specific Readmission Risk Stratification in a Predominantly Black Statewide Cohort Using Machine Learning: Development of Subtype-Specific Models for Heart Failure, Acute Myocardial Infarction, Atrial Fibrillation/Flutter, and Hypertensive Heart Disease

**DOI:** 10.64898/2026.03.08.26347901

**Authors:** Ismail El Moudden, Michael Bittner, Sunita Dodani

**Author notes:** Correspondence: Ismail El Moudden, MSc, PhD, PStat^®^, Macon & Joan Brock Virginia Health Sciences at Old Dominion University, 855 W Brambleton Ave, Williams Hall, Suite A125, Norfolk, VA 23510-1005.

## Abstract

**Background:** Cardiovascular disease (CVD) readmissions impose substantial clinical and economic burden. Machine learning (ML) may improve risk stratification, yet most predictive models aggregate CVD subtypes into a single outcome and underrepresent Black populations.

**Methods:** Using Virginia Health Information database records (2010 to 2020), we analyzed 157,791 discharge records from 123,272 unique patients (96.6% Black) to develop condition-specific 30-day readmission models for heart failure (HF; n=91,752), acute myocardial infarction (AMI; n=34,497), atrial fibrillation/flutter (AF/AFL; n=18,424), and hypertensive heart disease (HHD; n=13,118). Four algorithms (XGBoost, LightGBM, Random Forest, Elastic Net) plus a Super Learner ensemble were trained on patient-grouped 70/30 splits with and without Synthetic Minority Oversampling Technique balancing. Models incorporated validated clinical indices (LACE, Charlson, Elixhauser) and administrative social determinants of health proxies.

**Results:** The overall 30-day readmission rate was 18.9%. Best area under the receiver operating characteristic curve (AUC) values by condition were HF 0.708 (95% CI, 0.701 to 0.716), AMI 0.706 (95% CI, 0.691 to 0.721), AF/AFL 0.732 (95% CI, 0.715 to 0.750), and HHD 0.758 (95% CI, 0.735 to 0.777). XGBoost was the top-performing algorithm for three of four subtypes. The LACE Index, Charlson Comorbidity Index, and insurance type were consistently the strongest predictors. Algorithm-native, aggregated, and SHAP-based importance measures converged on these key features.

**Conclusions:** In this largest-to-date, predominantly Black statewide cohort, condition-specific ML models achieved moderate-to-high discrimination for HF, AMI, AF/AFL, and HHD. Key clinical indices and administrative social determinants proxies emerged as dominant predictors, highlighting modifiable targets and high-risk subgroups. These findings support the development of precision, equity-informed readmission interventions and provide a scalable framework for deploying ML-driven decision support in safety-net and minority-serving healthcare systems.

**WHAT IS KNOWN:** • Machine learning models for cardiovascular readmission prediction have largely aggregated disease subtypes and underrepresented Black populations.

• Most existing studies lack head-to-head algorithm comparisons within racially concentrated cohorts and omit social determinants of health proxies.

**WHAT THE STUDY ADDS:** • Condition-specific models for four cardiovascular subtypes achieved moderate-to-high discrimination (AUC 0.690 to 0.706) in the largest machine learning-based analysis of a predominantly Black statewide cohort.

• Validated clinical indices (LACE, Charlson) and insurance type consistently emerged as dominant predictors, identifying modifiable targets for equity-informed intervention.

• The scalable, administrative-data-only framework supports deployment of subtype-specific readmission decision support in safety-net and minority-serving health systems.

## Introduction

Cardiovascular disease (CVD) remains the leading cause of mortality in the United States, accounting for approximately 931,578 deaths annually and $252 billion in direct medical costs.^1,2^ Heart failure (HF), acute myocardial infarction (AMI), atrial fibrillation and flutter (AF/AFL), and hypertensive heart disease (HHD) are the principal drivers of cardiovascular hospitalization and 30-day readmission.^1–3^ Reported 30-day readmission rates range from 18% to 25% for HF,^3–5^ 12% to 17% for AMI,^6,7^ and 10% to 18% for AF/AFL,^8,9^ while HHD drives acute care utilization with disproportionately elevated rates in minority populations.^2,10^

Centers for Medicare & Medicaid Services (CMS) value-based penalty programs, including the Hospital Readmissions Reduction Program (HRRP) and the Hospital-Acquired Condition Reduction Program (HACRP), have produced modest improvements in quality metrics, yet safety-net hospitals serving predominantly Black and low-income patients bear a disproportionate share of financial penalties.^11–13^ Racial differences in AMI readmission persisted after HRRP and were attributable to patient-level rather than hospital-level factors.^12^ Black patients hospitalized for HF have 3.9% to 6.8% higher composite readmission and mortality rates than White patients across deprivation strata, even after covariate adjustment.^14^ Historical structural factors such as residential redlining compound these effects at the population level rather than at individual hospitals, pointing to the need for upstream interventions.^15,16^

Machine learning (ML) handles high-dimensional data and non-linear relationships more flexibly than logistic regression; AUC values from 0.51 to 0.93 have been reported across heart failure readmission studies.^17–20^ Most of these studies lacked external validation, rarely assessed calibration, and relied on racially heterogeneous populations unlikely to generalize to racially concentrated cohorts.^20^ Integrating social determinants of health (SDOH) into ML models has improved prediction in other cardiovascular contexts (AUC 0.694 to 0.823 for stroke), but application to readmission in predominantly minority populations has not been studied.^21,22^ Despite this growing body of work, ML models for CVD readmission have rarely been developed in predominantly minority populations,^14,16,19,20^ head-to-head algorithm comparisons across distinct cardiovascular conditions using standardized feature sets remain sparse,^5,19,20^ and no prior study has integrated validated clinical indices with administrative SDOH proxy data into condition-specific models within racially concentrated cohorts.^14,21,22^ We compared four ML algorithms (XGBoost, LightGBM, Random Forest, Elastic Net) and a Super Learner stacked ensemble across four conditions (HF, AMI, AF/AFL, HHD) in 157,791 discharge records from 123,272 unique patients (96.6% Black/African American) using an algorithm benchmarking design (4 base algorithms × 4 conditions × 2 dataset configurations plus 8 ensemble models, 40 total; TRIPOD Type 1a: development)^23^ with validated clinical indices and consensus-selected features. CVD conditions were classified using AHA-aligned ICD-9/ICD-10 hierarchies. The study design follows TRIPOD guidelines for model development studies (Type 1a),^23^ with the crossed design extending the framework to support simultaneous algorithm comparison within each condition. Similar crossed designs have been used in cardiovascular ML benchmarking.^5,18–20^ This study was conducted to evaluate the performance of machine learning models across different cardiovascular disease (CVD) subtypes in a minority population and to identify the relative importance of validated clinical indices and administrative social determinants of health (SDOH) proxies, particularly insurance status and comorbidity burden, on condition-specific risk prediction. We hypothesize that (1) ML model performance will differ across CVD subtypes, with condition-specific risk architectures requiring tailored modeling, and (2) validated clinical indices and administrative SDOH proxies particularly insurance status and comorbidity burden, will be significant predictors, with their relative importance varying by CVD subtype in this minority population. HHD was included as a standalone category because it disproportionately affects Black populations, who constitute 96.6% of our cohort, and is a leading contributor to heart failure progression, yet it has been largely absent from the readmission prediction literature as a separately modeled condition.

## Methods

### Data Availability Statement

The data that support the findings of this study are available from the Virginia Health Information (VHI) database. Restrictions apply to the availability of these data, which were used under a data use agreement for the current study. Data may be available from VHI (https://www.vhi.org) upon reasonable request and with permission. The analytic code is available in a private GitHub repository (https://github.com/isamil/ML-CVD-Readmission-in-Black-Cohort); access will be granted to reviewers upon request and the repository will be made public upon publication. Dr. El Moudden had full access to all the data in the study and takes responsibility for its integrity and the data analysis.

### Study Design and Data Source

We conducted a retrospective cohort study using de-identified inpatient discharge records from the Virginia Health Information (VHI) statewide all-payer database (January 2010 through December 2020).^24^ VHI captures demographic, clinical, financial, and administrative data from all acute-care hospitalizations in Virginia (approximately 9.24 million total discharges). The study objective was to develop and compare ML models for 30-day all-cause readmission across four CVD subtypes in a predominantly minority population. Cohort flow is illustrated in **Figure 1**.

**Figure 1.**
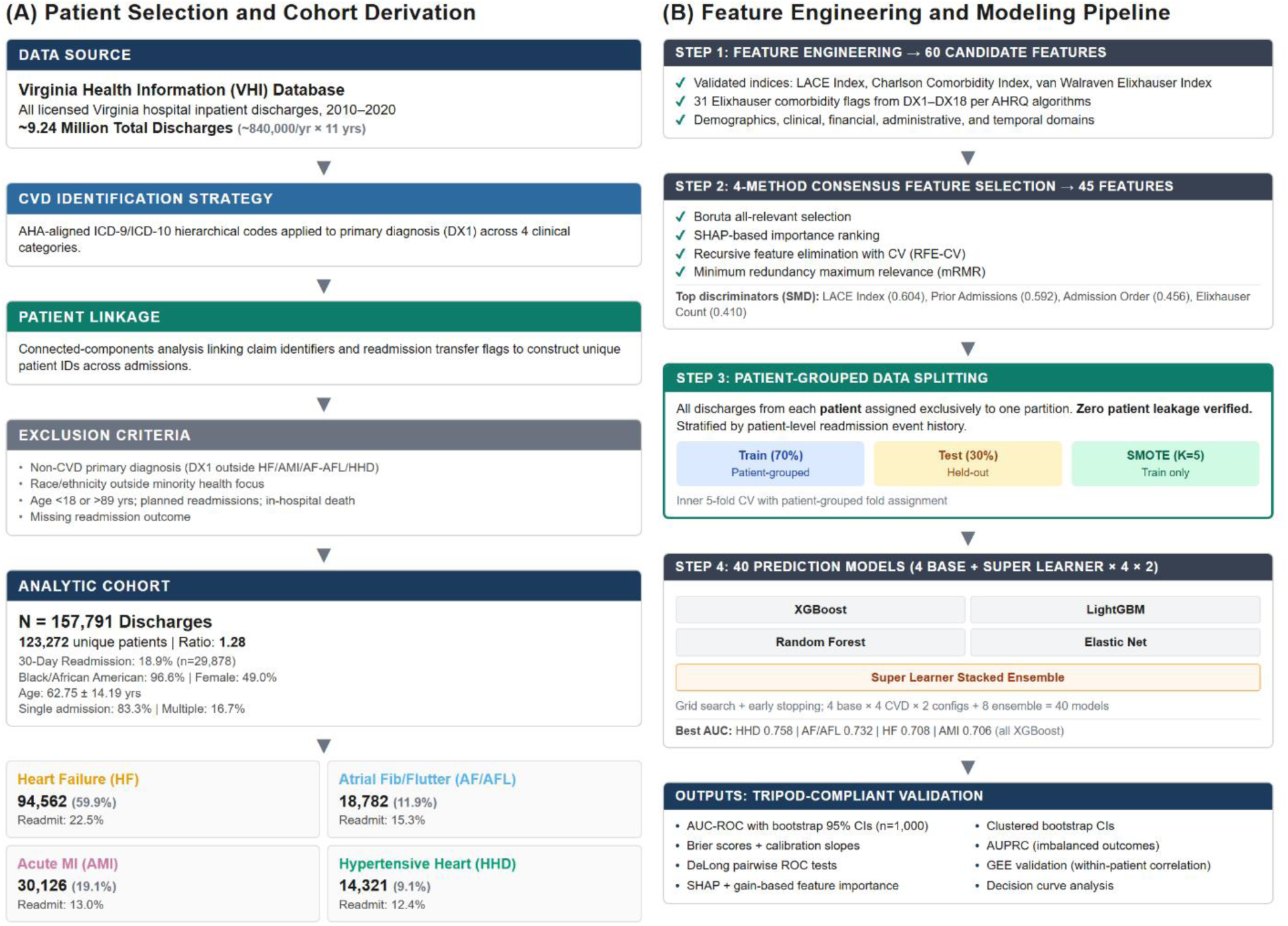
Study Design and Cohort Flow. CONSORT-style diagram showing cohort derivation from the VHI statewide database (157,791 discharges from 123,272 patients), patient-grouped 70/30 train-test split, four CVD category subsets, balanced and unbalanced configurations, 4 base algorithms plus Super Learner ensemble, totaling 40 models. Abbreviations: AF/AFL, atrial fibrillation/flutter; AMI, acute myocardial infarction; CVD, cardiovascular disease; HF, heart failure; HHD, hypertensive heart disease; ML, machine learning; SMOTE, Synthetic Minority Oversampling Technique; VHI, Virginia Health Information.

### CVD Identification

CVD patients were identified from the primary diagnosis field using AHA guideline-aligned hierarchical classification. Four categories were defined: HF, AMI, AF/AFL, and HHD. Dual-era diagnostic codes (ICD-9 and ICD-10) were mapped to each category; complete code lists are available in Supplementary **Table S1**.

### Exclusion Criteria

We excluded records with non-CVD primary diagnoses, race/ethnicity other than Black/African American or Hispanic/Latino (per the grant’s minority health focus), age < 18 or >89 years, planned readmissions, in-hospital death, or missing readmission outcome. After exclusions, 157,791 discharge records from 123,272 unique patients comprised the analytic cohort; no records were excluded for missing patient identifiers. Baseline characteristics are presented in Table 1. The analytic cohort was 96.6% Black/African American and 3.4% Hispanic/Latino, consistent with the grant’s minority health focus. Given the small Hispanic subgroup, no separate models were estimated for this population; however, model performance was confirmed to be comparable when restricted to Black patients alone (results available upon request).

**Table 1.** Baseline Demographics and Clinical Characteristics by Readmission Status. (N = 157,791 Discharge Records, 123,272 Unique patients, 1.28 Discharge-to-patient ratio, 18.9% Readmission rate)

| Characteristic | Overall<br>(N = 157,791) | No Readmission<br>(n = 127,913) | Readmission<br>(n = 29,878) | SMD |
| --- | --- | --- | --- | --- |
| **Demographics and Encounter Characteristics** |  |  |  |  |
| Age, years, mean (SD) | 62.75 (14.19) | 62.55 (14.17) | 63.60 (14.23) | 0.074 |
| Female sex, % | 49.0 | 49.0 | 51.0 | 0.036 |
| Length of stay, days, mean (SD) | 4.99 (6.01) | 4.81 (5.98) | 5.73 (6.08) | 0.151 |
| Total diagnoses, mean (SD) | 14.41 (4.14) | 14.13 (4.24) | 15.60 (3.44) | **0.381** |
| Total procedures, mean (SD) | 1.72 (1.96) | 1.76 (1.99) | 1.56 (1.85) | 0.100 |
| Total charges (\$), mean (SD) | 47,910 (88,915) | 47,727 (88,549) | 48,692<br>(90,459) | 0.011 |
| Admission Order, mean (SD) | 1.49 (1.59) | 1.31 (1.04) | 2.28 (2.82) | **0.456** |
| N Admissions per Patient, mean (SD) | 1.99 (2.63) | 1.60 (1.68) | 3.65 (4.59) | **0.592** |
| **Validated Clinical Indices** |  |  |  |  |
| LACE Index (0–19), mean (SD) | 9.67 (3.02) | 9.34 (2.96) | 11.09 (2.86) | **0.604** |
| Charlson Comorbidity Index, mean (SD) | 3.15 (2.05) | 2.99 (2.04) | 3.81 (1.99) | **0.405** |
| Age-Adjusted Charlson, mean (SD) | 5.04 (2.65) | 4.87 (2.65) | 5.78 (2.48) | **0.357** |
| van Walraven Elixhauser Index, mean (SD) | 11.99 (8.74) | 11.34 (8.70) | 14.74 (8.37) | **0.398** |
| Elixhauser Comorbidity Count, mean (SD) | 4.63 (2.00) | 4.49 (2.00) | 5.27 (1.85) | **0.410** |
| CVD Severity Score, mean (SD) | 3.49 (0.81) | 3.45 (0.83) | 3.67 (0.65) | **0.294** |
| CVD Mortality Risk, mean (SD) | 2.65 (0.90) | 2.62 (0.92) | 2.79 (0.83) | 0.199 |
| Total CVD Conditions, mean (SD) | 4.57 (1.94) | 4.50 (1.94) | 4.88 (1.91) | 0.195 |
| **LACE Risk Category, n (%)** |  |  |  | **0.502** |
| Low | 6,188 (3.9) | 5,827 (4.6) | 361 (1.2) |  |
| Moderate | 67,567 (42.8) | 59,606 (46.6) | 7,961 (26.6) |  |
| High | 84,036 (53.3) | 62,480 (48.8) | 21,556 (72.1) |  |
| **Race/Ethnicity, n (%)** |  |  |  | 0.066 |
| Black | 152,492 (96.6) | 123,344 (96.4) | 29,148 (97.6) |  |
| Hispanic | 5,299 (3.4) | 4,569 (3.6) | 730 (2.4) |  |
| **Insurance Type, n (%)** |  |  |  | **0.342** |
| Commercial | 33,341 (21.1) | 29,266 (22.9) | 4,075 (13.6) |  |
| Medicaid | 16,868 (10.7) | 12,881 (10.1) | 3,987 (13.3) |  |
| Medicare | 92,250 (58.5) | 72,024 (56.3) | 20,226 (67.7) |  |
| Self-Pay/Uninsured | 15,332 (9.7) | 13,742 (10.7) | 1,590 (5.3) |  |
| **Primary CVD Category, n (%)** |  |  |  | **0.302** |
| Acute Myocardial Infarction | 30,126 (19.1) | 26,207 (20.5) | 3,919 (13.1) |  |
| Atrial Fibrillation/Flutter | 18,782 (11.9) | 15,902 (12.4) | 2,880 (9.6) |  |
| Heart Failure | 94,562 (59.9) | 73,253 (57.3) | 21,309 (71.3) |  |
| Hypertensive Heart Disease | 14,321 (9.1) | 12,551 (9.8) | 1,770 (5.9) |  |
| **Teaching Hospital Status, n (%)** |  |  |  | 0.083 |
| ACGME | 22,062 (14.0) | 18,105 (14.2) | 3,957 (13.3) |  |
| Council for GME | 13,626 (8.6) | 11,029 (8.6) | 2,597 (8.7) |  |
| Council of Teaching | 9,082 (5.8) | 7,006 (5.5) | 2,076 (7.0) |  |
| Council of Teaching Hospitals | 19,534 (12.4) | 15,447 (12.1) | 4,087 (13.7) |  |
| None | 93,325 (59.2) | 76,190 (59.6) | 17,135 (57.4) |  |
| **Hospital Size (beds), n (%)** |  |  |  | 0.069 |
| Small | 17,088 (10.8) | 13,823 (10.8) | 3,265 (10.9) |  |
| Medium | 60,458 (38.3) | 49,587 (38.8) | 10,871 (36.4) |  |
| Large | 47,702 (30.2) | 38,757 (30.3) | 8,945 (29.9) |  |
| Major | 32,543 (20.6) | 25,746 (20.1) | 6,797 (22.7) |  |
| **Hospital Ownership, n (%)** |  |  |  | 0.028 |
| Not-for-Profit | 125,919 (79.8) | 102,345 (80.0) | 23,574 (78.9) |  |
| Proprietary | 31,871 (20.2) | 25,567 (20.0) | 6,304 (21.1) |  |
| **Elixhauser Comorbidities (prevalence), %** |  |  |  |  |
| Congestive Heart Failure | 68 | 66 | 78 | **0.268** |
| Renal Failure | 45 | 42 | 59 | **0.325** |
| Cardiac Arrhythmia | 37 | 36 | 42 | 0.132 |
| Chronic Pulmonary Disease | 35 | 33 | 43 | 0.199 |
| Fluid & Electrolyte Disorder | 32 | 30 | 36 | 0.120 |
| Hypertension, Complicated | 31 | 29 | 35 | 0.124 |
| Obesity | 28 | 29 | 25 | 0.085 |
| Hypertension, Uncomplicated | 25 | 27 | 18 | **0.201** |
| Diabetes, Uncomplicated | 25 | 25 | 26 | 0.018 |
| Diabetes with Complications | 25 | 24 | 30 | 0.142 |
| Pulmonary Circulation Disorder | 19 | 18 | 23 | 0.135 |
| Valvular Disease | 18 | 18 | 20 | 0.067 |
| Peripheral Vascular Disease | 11 | 11 | 13 | 0.079 |
| Depression | 8 | 7 | 10 | 0.079 |
| Hypothyroidism | 8 | 8 | 9 | 0.050 |
| Other Neurological Disorder | 6 | 6 | 8 | 0.090 |
| Drug Abuse | 6 | 6 | 7 | 0.055 |
| Deficiency Anemia | 6 | 6 | 7 | 0.047 |
| Alcohol Abuse | 5 | 5 | 5 | 0.008 |
| Coagulopathy | 5 | 5 | 7 | 0.059 |
| Weight Loss | 4 | 4 | 6 | 0.098 |
| Liver Disease | 4 | 4 | 6 | 0.087 |
| Rheumatoid Arthritis/Collagen | 3 | 3 | 4 | 0.049 |
| Psychoses | 2 | 2 | 2 | 0.057 |
| Solid Tumor w/o Metastasis | 2 | 2 | 3 | 0.063 |
| Metastatic Cancer | 1 | 1 | 1 | 0.061 |
| Lymphoma | 1 | 1 | 1 | 0.044 |
| Blood Loss Anemia | 1 | 1 | 1 | 0.032 |
| HIV/AIDS | 1 | 0 | 1 | 0.033 |
| Peptic Ulcer Disease | 0 | 0 | 1 | 0.013 |
| Paralysis | 1 | 1 | 1 | 0.013 |

Abbreviations: ACGME, Accreditation Council for Graduate Medical Education; CVD, cardiovascular disease; GME, Graduate Medical Education; LACE, Length of stay + Acuity + Comorbidities + Emergency visits; SD, standard deviation; SMD, standardized mean difference.
Total discharge records: N = 157,791 from 123,272 unique patients (discharge-to-patient ratio = 1.28).
Values are mean (SD) for continuous variables and n (%) or prevalence (%) for categorical variables. SMD ≥ 0.20 (bolded) indicates a meaningful difference between groups. For categorical variables with >2 levels, a single overall SMD is reported on the section header row.
Validated indices computed from discharge data: LACE Index (van Walraven et al., 2010), Charlson Comorbidity Index (Charlson et al., 1987; Quan et al., 2011), van Walraven Elixhauser Index (van Walraven et al., 2009). All 31 Elixhauser comorbidity flags shown with individual SMDs.

### Unit of Analysis and Patient Grouping

The unit of analysis was the discharge record, matching the HRRP penalty structure.^11,12^ Because patients could contribute multiple admissions, all data partitions were grouped at the patient level to prevent information leakage. A 70/30 train-test split assigned all discharges from a given patient exclusively to one partition.

Hyperparameter tuning used inner 5-fold patient-grouped cross-validation. Zero patient overlap was programmatically verified. Prior large-scale administrative readmission studies have used the same grouping strategy.^4,6,18,20^ To evaluate whether discrimination differed by admission history, test-set performance was stratified by admission order (first admission vs. subsequent admissions) for each best-performing model (**Supplementary Table S6**).

### Outcome Definition

The primary outcome was 30-day all-cause readmission to any Virginia acute-care hospital, identified through longitudinal patient linkage using Readmissions and Transfers (RATs) supplemental files.

### Feature Engineering

Raw administrative data were supplemented with validated clinical indices pre-computed during data extraction: the LACE Index (van Walraven et al., 2010),^32^ Charlson Comorbidity Index (Charlson 1987; Quan 2005),^33,34^ Age-Adjusted Charlson Index, van Walraven Elixhauser Index (van Walraven et al., 2009),^35^ and Elixhauser Comorbidity Count. All 31 AHRQ Elixhauser comorbidity flags were pre-computed from secondary diagnosis fields (DX2–DX18) using published ICD-9/ICD-10 mappings.^25^ Additional features captured hospital characteristics (teaching status, ownership, bed size), admission characteristics (length of stay, total diagnoses, total procedures), and patient history (admission order [sequential index of each discharge], admissions per patient [cumulative count]). Complete feature specifications appear in Supplementary **Table S1**.

The study period (January 2010 through December 2020) spans the October 1, 2015 transition from ICD-9-CM to ICD-10-CM coding. Primary CVD diagnosis classification used published dual-era diagnostic mappings (**Supplementary Table S1**). However, ICD-10’s substantially greater coding granularity may have inflated comorbidity counts and altered Elixhauser flag prevalence in the post-transition period, as previously documented in administrative database studies. Admission Year was included as a temporal feature (selected by 3 of 4 feature selection methods), which may partially absorb coding era effects. A descriptive comparison of key comorbidity features across coding eras is provided in **Supplementary Table S10**.

### Feature Selection

From 60 candidate features after technical filtering (removal of zero-variance, near-zero-variance features with >95% single value, and one member of pairs with |r| > 0.90), four independent selection methods were applied to the training set: (a) Boruta all-relevant selection,^36^ (b) SHAP-based importance from a reference XGBoost model, (c) recursive feature elimination with patient-grouped cross-validation, and (d) minimum redundancy maximum relevance (mRMR).^37^ Features endorsed by at least two of four methods were retained, producing 45 consensus-selected features applied uniformly across all CVD categories and configurations. This data-driven consensus approach replaced the univariate correlation ranking used in earlier analyses, removing the arbitrary top-k cap so that retained features received converging evidence from multiple methodologically distinct selection criteria (Supplementary **Table S1**, **Figure S1**).

### Machine Learning Model Development

Four base algorithms were evaluated: XGBoost, LightGBM, Random Forest (ranger), Elastic Net (glmnet). Each was trained independently for each CVD category under two configurations (unbalanced and SMOTE-balanced), for a total of 32 base models. A Super Learner stacked ensemble combined out-of-fold predictions from all base learners through a non-negatively constrained logistic regression meta-learner,40 adding 8 ensemble models (one per CVD category per dataset configuration) for a total of 40 models (4 base × 4 categories × 2 configurations + 8 ensemble; **Figure 1**). All metrics were evaluated on the held-out test set, held out from training, tuning, and SMOTE augmentation. Balanced training sets used SMOTE with K = 5 nearest neighbors applied exclusively to the training partition. For unbalanced datasets, class weights were assigned through algorithm-native methods (scale_pos_weight, inverse-class-frequency weighting, or native logistic loss). All algorithms underwent grid search with inner 5-fold patient-grouped cross-validation maximizing AUC. XGBoost and LightGBM each searched 27 configurations with early stopping at 15 rounds; Random Forest searched 18 configurations selected by out-of-bag error; Elastic Net optimized 11 alpha values with lambda selected by the one-standard-error rule. The term “ensemble” in the title refers to the Super Learner stacked ensemble approach, which combines out-of-fold predictions from all base learners through a non-negatively constrained logistic regression meta-learner.

### Performance Assessment and Validation

Model performance followed TRIPOD guidelines^23^ on the 30% held-out test set. Discrimination was assessed by AUC with 95% bootstrap Cis (1,000 iterations, percentile method) and area under the precision-recall curve (AUPRC), which is more informative than AUC-ROC for imbalanced outcomes. The optimal threshold was determined by maximizing the Youden index, at which sensitivity, specificity, positive and negative predictive values, F1 score, balanced accuracy, and Matthews correlation coefficient were computed. Clinical utility was classified as Excellent (≥0.80), High (≥0.75), Moderate (≥0.70), or Limited (<0.70). Calibration was assessed using Brier scores, calibration slopes, calibration-in-the-large (CITL), and expected-to-observed (E/O) ratio per TRIPOD guidelines (**Figure 4**). ROC curves are presented in **Figure 2** and the AUC performance heatmap in **Figure 3**. Pairwise algorithm comparisons used DeLong tests within the best-performing dataset configuration per CVD category (**Supplementary Table S3**).

**Figure 2.**
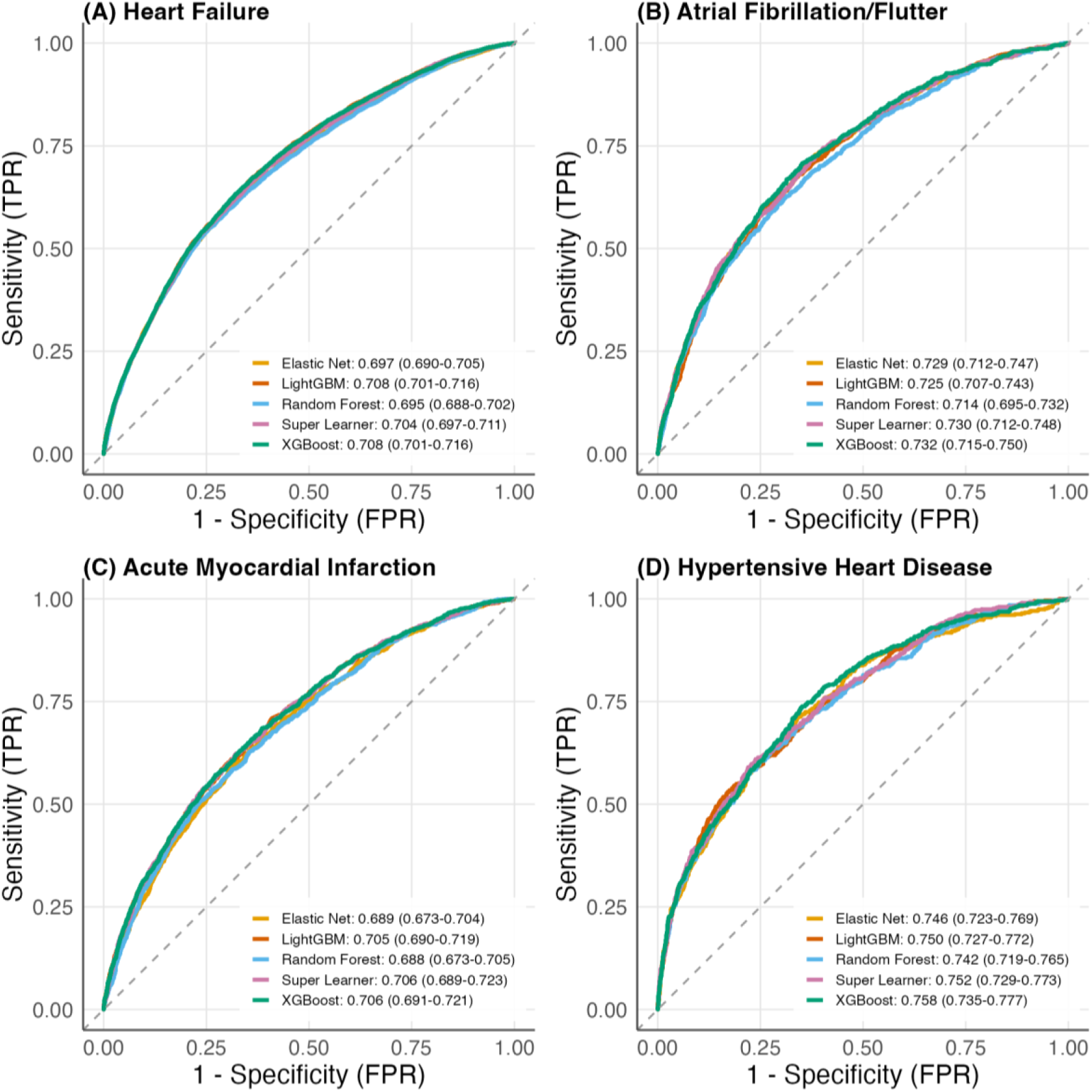
Receiver Operating Characteristic Curves by CVD Category. ROC curves for all algorithms across four CVD categories (HF, AMI, AF/AFL, HHD) under balanced and unbalanced configurations. AUC values reported in legends. Abbreviations: AF/AFL, atrial fibrillation/flutter; AMI, acute myocardial infarction; AUC, area under the receiver operating characteristic curve; CI, confidence interval; CVD, cardiovascular disease; FPR, false positive rate; HF, heart failure; HHD, hypertensive heart disease; ROC, receiver operating characteristic; TPR, true positive rate.

**Figure 3.**
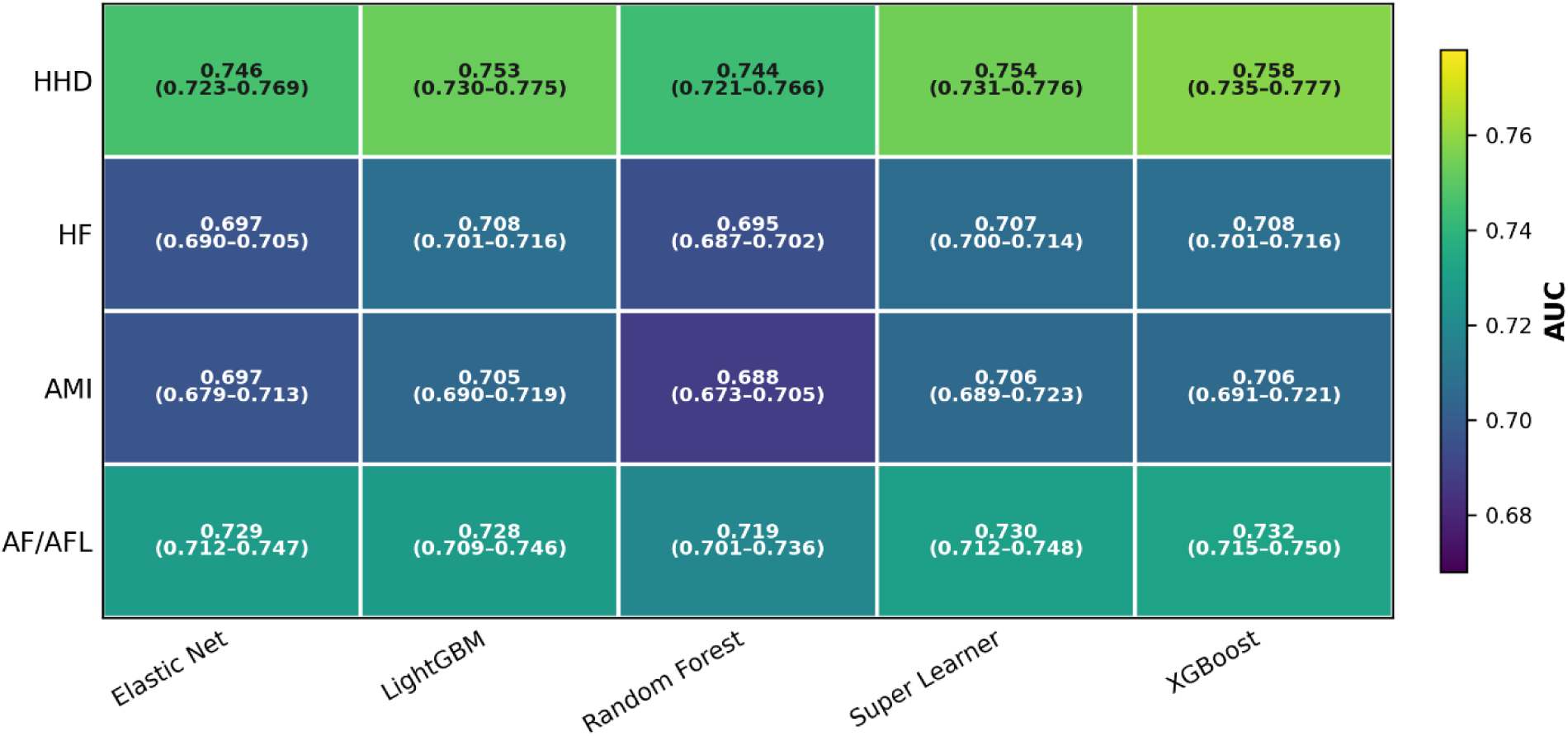
AUC Performance Heatmap. Heatmap displaying AUC values for all 40 models (5 algorithms × 4 CVD categories × 2 configurations). Color scale indicates utility classification. Abbreviations: AF/AFL, atrial fibrillation/flutter; AMI, acute myocardial infarction; AUC, area under the receiver operating characteristic curve; CI, confidence interval; CVD, cardiovascular disease; HF, heart failure; HHD, hypertensive heart disease.

**Figure 4.**
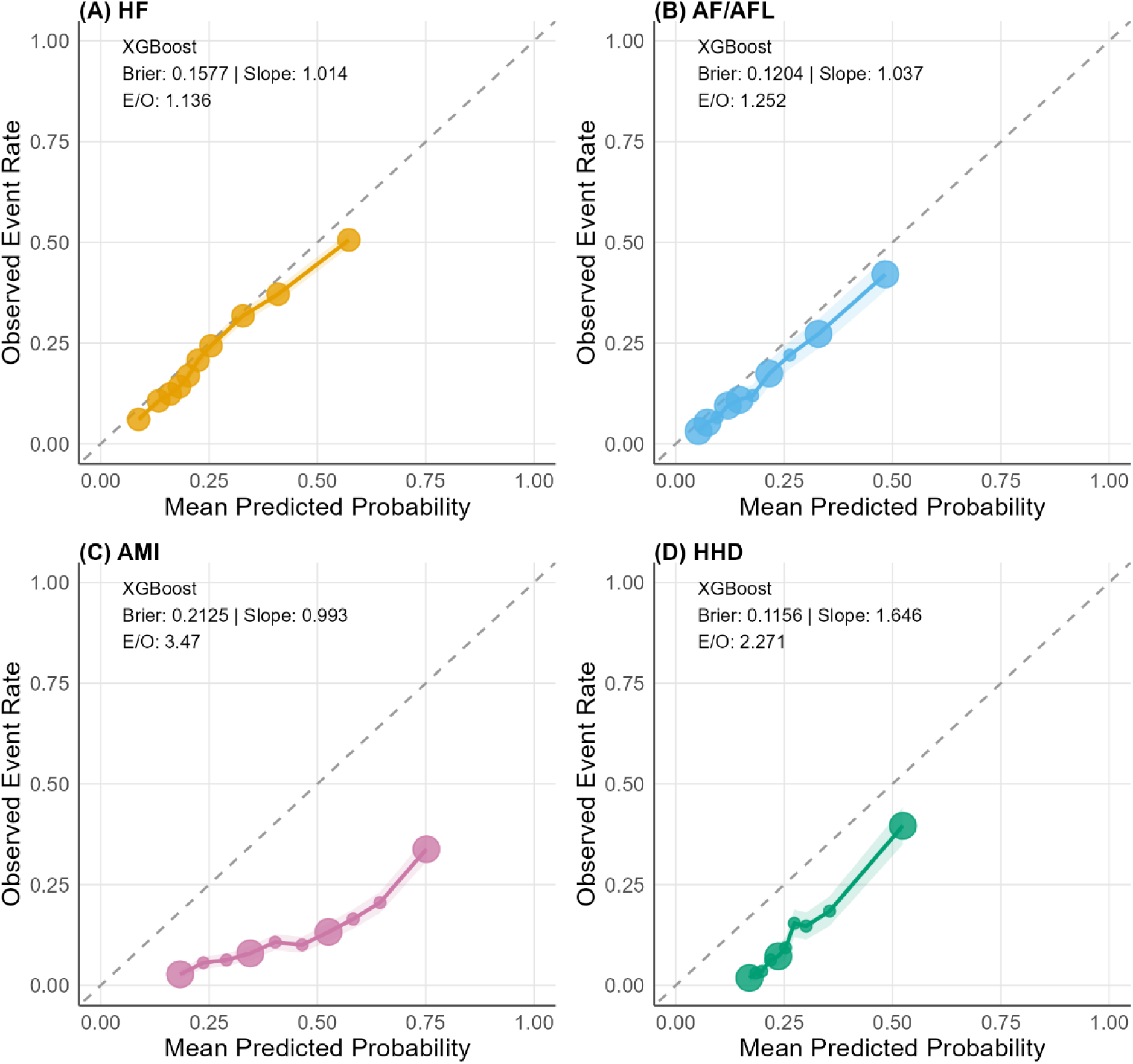
Calibration Plots for Best-Performing Models. Calibration curves for the best model per CVD category with calibration slope, intercept, CITL, and E/O ratio annotated. Abbreviations: AF/AFL, atrial fibrillation/flutter; AMI, acute myocardial infarction; CI, confidence interval; CVD, cardiovascular disease; E/O, expected-to-observed ratio; HF, heart failure; HHD, hypertensive heart disease.

To assess residual within-patient correlation, we fitted GEE models with exchangeable working correlation and sandwich standard errors on multi-admission patient subsets within each test partition. Patient-resampled clustered bootstrap CIs (1,000 iterations) were also computed.

Results appear in Supplementary Tables S4–S5.

### Feature Importance Analysis

Feature importance was quantified using algorithm-native methods (gain-based for XGBoost, LightGBM; impurity-based for Random Forest; absolute standardized coefficients for Elastic Net), normalized to [0, 1] within each model. Three complementary perspectives are reported: condition-specific importance from each best-performing model (**Figure 5**), aggregated importance averaged across all 32 base models requiring appearance in at least three (**Supplementary Tables S7 and S8**), and SHapley Additive exPlanations (SHAP)-based importance providing directional, additive decompositions grounded in cooperative game theory (**Figure 6**, **Supplementary Figures S3–S4**). Algorithm-native importance served as the primary measure; SHAP analysis provides supplementary mechanistic insight.

**Figure 5.**
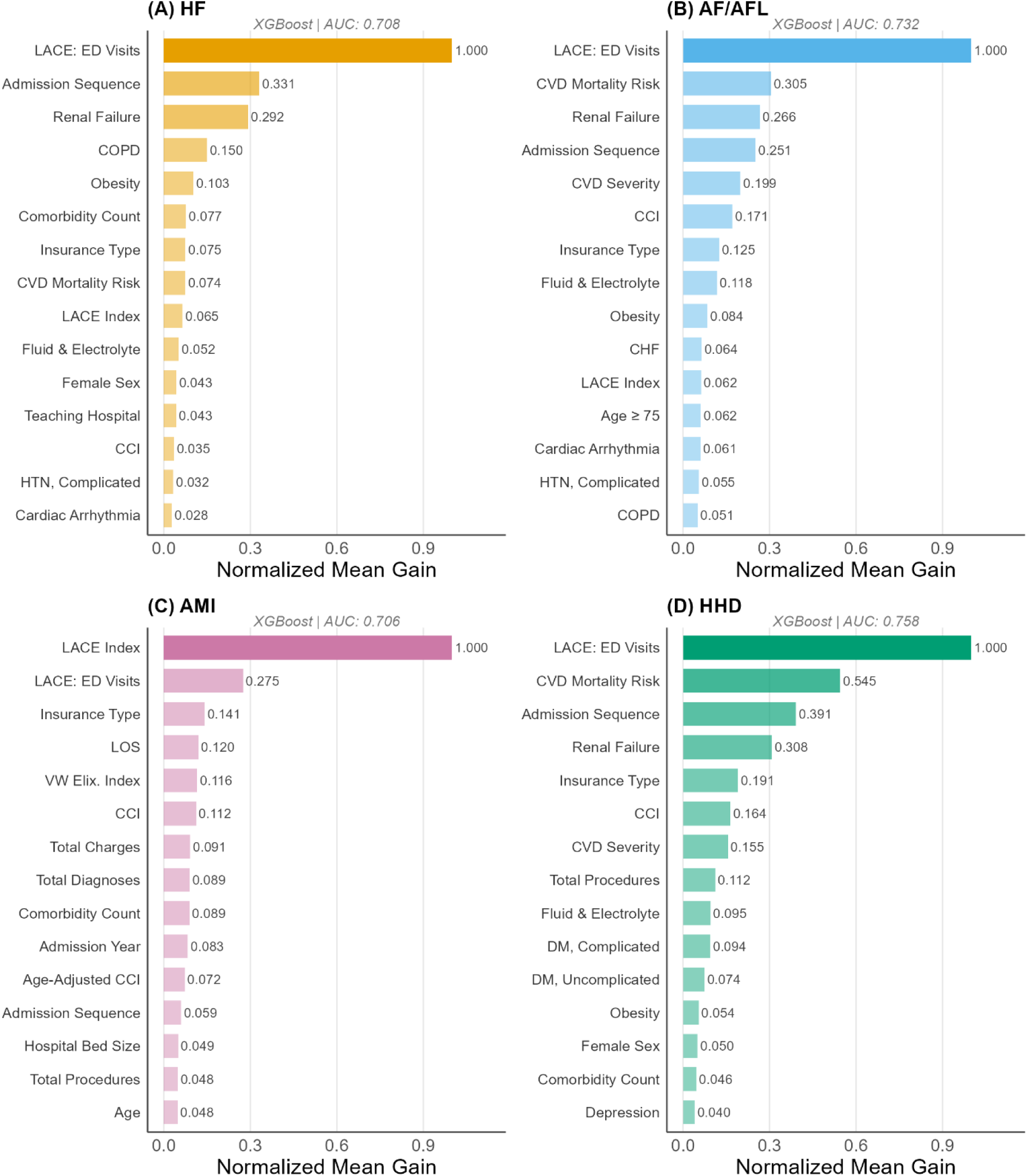
Feature Importance: Top 15 Predictors by Mean Gain. Horizontal bar chart showing top 15 features by mean gain-based importance across best-performing models, colored by feature domain. Abbreviations: AF/AFL, atrial fibrillation/flutter; AMI, acute myocardial infarction; AUC, area under the receiver operating characteristic curve; CVD, cardiovascular disease; HF, heart failure; HHD, hypertensive heart disease.

**Figure 6.**
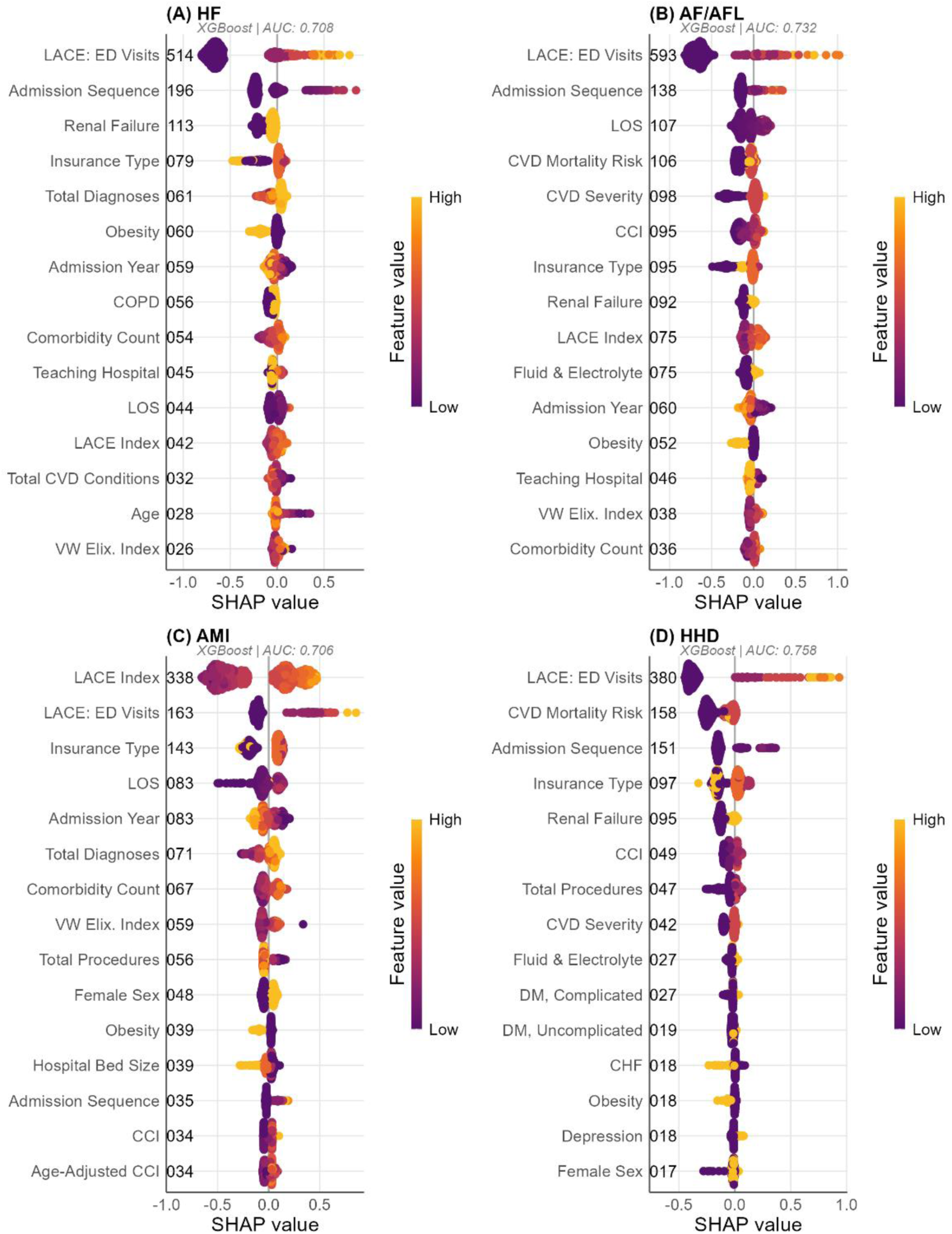
SHAP Beeswarm Summary Plots. SHAP summary plots for the best-performing model per CVD category showing top 20 features with direction and magnitude of effects on predicted readmission risk. Abbreviations: AF/AFL, atrial fibrillation/flutter; AMI, acute myocardial infarction; AUC, area under the receiver operating characteristic curve; CVD, cardiovascular disease; HF, heart failure; HHD, hypertensive heart disease; SHAP, SHapley Additive exPlanations.

Post-hoc SHAP analysis was performed on each condition’s best-performing model (**Table 2**) using shapviz in R. Exact TreeSHAP values were computed for tree-based models (XGBoost, LightGBM) via native contribution output. SHAP values were computed on a random subsample of 500 test observations per condition (set.seed(42)). Three visualizations were generated: beeswarm summary plots (**Figure 6**), waterfall plots for representative true positive and true negative cases (**Supplementary Figure S4**), and dependence plots for continuous predictors (**Supplementary Figure S3**). SHAP analysis was performed post-hoc and did not influence model selection or hyperparameter tuning. SHAP values were computed on a random subsample of 500 test observations per CVD category due to computational cost. The subsample was verified for representativeness by comparing event rates and distributions of the top 5 predictors (LACE Index, Charlson Comorbidity Index, Length of Stay, Elixhauser Comorbidity Count, and Admission Order) between the subsample and the full test set, confirming no meaningful divergence (all standardized mean differences < 0.10).

**Table 2.**
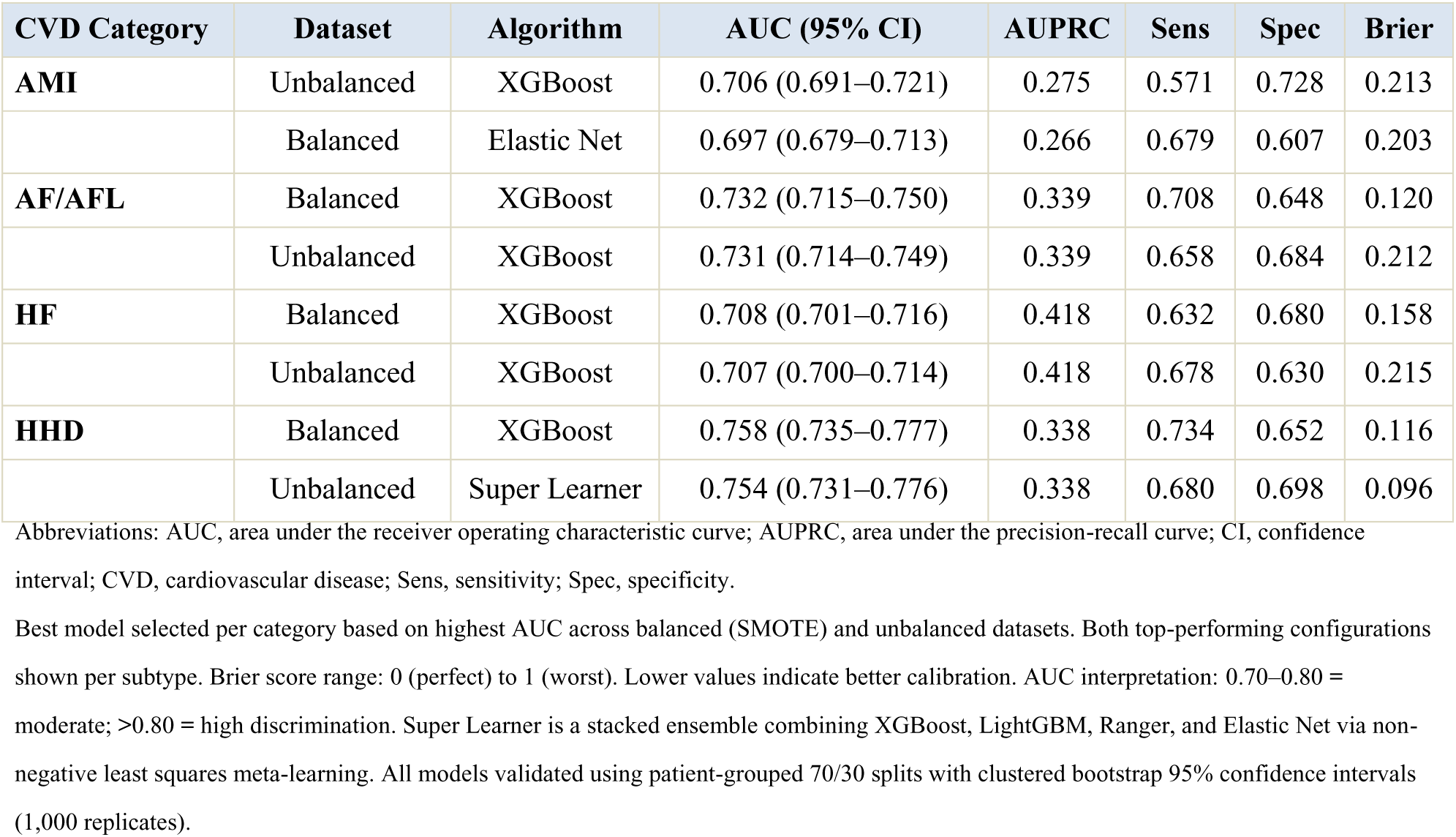
Best-Performing Models by CVD Category.

| CVD Category | Dataset | Algorithm | AUC (95% CI) | AUPRC | Sens | Spec | Brier |
| --- | --- | --- | --- | --- | --- | --- | --- |
| AMI | Unbalanced | XGBoost | 0.706 (0.691–0.721) | 0.275 | 0.571 | 0.728 | 0.213 |
|  | Balanced | Elastic Net | 0.697 (0.679–0.713) | 0.266 | 0.679 | 0.607 | 0.203 |
| AF/AFL | Balanced | XGBoost | 0.732 (0.715–0.750) | 0.339 | 0.708 | 0.648 | 0.120 |
|  | Unbalanced | XGBoost | 0.731 (0.714–0.749) | 0.339 | 0.658 | 0.684 | 0.212 |
| HF | Balanced | XGBoost | 0.708 (0.701–0.716) | 0.418 | 0.632 | 0.680 | 0.158 |
|  | Unbalanced | XGBoost | 0.707 (0.700–0.714) | 0.418 | 0.678 | 0.630 | 0.215 |
| HHD | Balanced | XGBoost | 0.758 (0.735–0.777) | 0.338 | 0.734 | 0.652 | 0.116 |
|  | Unbalanced | Super Learner | 0.754 (0.731–0.776) | 0.338 | 0.680 | 0.698 | 0.096 |

Abbreviations: AUC, area under the receiver operating characteristic curve; AUPRC, area under the precision-recall curve; CI, confidence interval; CVD, cardiovascular disease; Sens, sensitivity; Spec, specificity.
Best model selected per category based on highest AUC across balanced (SMOTE) and unbalanced datasets. Both top-performing configurations shown per subtype. Brier score range: 0 (perfect) to 1 (worst). Lower values indicate better calibration. AUC interpretation: 0.70–0.80 = moderate; >0.80 = high discrimination. Super Learner is a stacked ensemble combining XGBoost, LightGBM, Ranger, and Elastic Net via non- negative least squares meta-learning. All models validated using patient-grouped 70/30 splits with clustered bootstrap 95% confidence intervals (1,000 replicates).

### Model Specification and Reproducibility

Tree ensemble models (XGBoost, LightGBM, Random Forest) and the Super Learner cannot be expressed as closed-form equations; their prediction logic is distributed across learned parameters. Serialized model objects containing all trained models, preprocessing parameters, hyperparameter configurations, selected features, and optimal thresholds were archived and are available from the corresponding author. A standalone prediction function accepting raw patient-level input and returning readmission probabilities was also archived for prospective deployment.

For Elastic Net, which yields interpretable coefficient vectors, **Supplementary Table S8** reports the full coefficient matrix (intercept plus 45 features) for each CVD category under both dataset configurations. These coefficients enable exact reproduction of predicted log-odds without access to the serialized model objects: log-odds(readmission) = β₀ + Σβⱼxⱼ, where β values are provided in Supplementary **Table S8** and λ was selected by the one-standard-error rule.

### Algorithmic Bias Assessment

Bias assessment included demographic parity across insurance types and age groups, equalized odds evaluation, calibration consistency analysis, and post-hoc auditing of differential prediction by insurance type.

### Statistical Analysis

All analyses were conducted in R version 4.4.2 using xgboost, lightgbm, ranger, glmnet, caret, smotefamily, pROC, geepack, Boruta, mRMRe, shapviz, and tableone. Baseline characteristics were compared using Welch’s t-tests and Pearson’s chi-squared tests. Effect sizes were reported as standardized mean differences (SMDs) computed via the tableone package, with SMD ≥0.10 considered indicative of meaningful imbalance. All 31 Elixhauser comorbidity flags were pre-computed during data extraction using published AHRQ ICD-9/ICD-10 mappings applied to secondary diagnosis fields.^25^ Two variables had missing values: Teaching Status (0.1%) and Hospital Ownership (<0.01%), handled by mode imputation. Four candidate features (APR-DRG Severity of Illness, APR-DRG Risk of Mortality, Admission Type, Discharge Disposition) were unavailable in the analytic dataset and excluded. A fixed random seed (set.seed(42)) was applied prior to all stochastic procedures.^26^ With a sample of 157,791 discharges, conventional significance tests (Welch’s t and Pearson’s chi-squared) have near-certainty of rejecting the null hypothesis for even trivially small differences. Therefore, while p-values are reported in **Table 1** for completeness, clinical meaningfulness of between-group differences was assessed using standardized mean differences (SMDs), with SMD > 0.10 indicating meaningful imbalance per established thresholds (Austin, 2009). No formal multiplicity adjustment was applied to model selection, consistent with standard practice in machine learning benchmarking studies. For each CVD category, the best-performing model was selected from 10 candidates (5 algorithms × 2 configurations); the optimistic bias inherent in selecting the maximum of multiple correlated estimates (“winner’s curse”) is acknowledged as a limitation.

### Ethical Considerations

The study was approved by the Eastern Virginia Medical School IRB (IRB #23-09-NH-0248) with waiver of informed consent for de-identified data analysis. All analyses used de-identified data with no attempt to re-identify patients.

## Results

### Cohort Characteristics

The analytic cohort comprised 157,791 discharge records from 123,272 unique patients identified through the VHI statewide database (discharge-to-patient ratio: 1.28), with 83.3% of patients contributing a single admission and 16.7% contributing two or more admissions. No records were excluded for missing outcome or patient identifiers. All ML model training and evaluation used this full cohort; cohort flow is detailed in **Figure 1**. Baseline demographics and clinical characteristics are presented in **Table 1**.

The population was 96.6% Black/African American and 49.0% female, with a mean age of 62.75±14.19 years. HF was the most common primary CVD category (59.9% of discharges), followed by AMI (19.1%), AF/AFL (11.9%), and HHD (9.1%). Readmitted patients had higher LACE Index scores (11.09±2.86 vs 9.34±2.96; SMD=0.604), more admissions per patient (3.65±4.59 vs 1.60±1.68; SMD=0.592), higher admission order (2.28±2.82 vs 1.31±1.04; SMD=0.456), greater Elixhauser comorbidity count (5.27±1.85 vs 4.49±2.00; SMD=0.410), higher Charlson Comorbidity Index (3.81±1.99 vs 2.99±2.04; SMD=0.405), higher van Walraven Elixhauser Index (14.74±8.37 vs 11.34±8.70; SMD=0.398), more total diagnoses (15.60±3.44 vs 14.13±4.24; SMD=0.381), and higher prevalence of renal failure (59% vs 42%; SMD=0.325). Insurance type showed substantial separation (SMD=0.342), with Medicare overrepresented among readmissions (67.7% vs 56.3%) and commercial insurance underrepresented (13.6% vs 22.9%).

### 30-Day Readmission Outcome

The overall 30-day all-cause readmission rate was 18.9% (n=29,878). Rates varied across CVD categories: HF 22.5%, AF/AFL 15.3%, AMI 13.0%, and HHD 12.4%. HF was disproportionately represented among readmissions (71.3% of readmitted discharges vs 57.3% of non-readmitted). Medicare beneficiaries accounted for 67.7% of readmissions compared with 56.3% of non-readmitted discharges (SMD=0.342), while self-pay/uninsured patients were underrepresented among readmissions (5.3% vs 10.7%). The LACE Risk Category showed the strongest categorical separation: 72.1% of readmitted patients were classified as high-risk compared with 48.8% of non-readmitted patients (SMD=0.502).

### Model Discrimination

From 60 candidate features after technical filtering, four independent selection methods yielded 45 consensus-selected features, applied uniformly across all CVD categories and dataset configurations. Forty prediction models were then trained by crossing four base algorithms plus a Super Learner ensemble with four CVD categories and two dataset configurations. All models used patient-grouped 70/30 splitting with zero patient leakage between partitions. Complete results for all 40 models appear in **Supplementary Table S2**; the best-performing models per category are summarized in **Table 2**.

Discrimination varied across CVD subtypes, with all four categories achieving moderate or high discrimination (AUC 0.706–0.758), though the highest-performing HHD model (AUC 0.758) exhibited calibration limitations (slope 1.479–1.646) requiring recalibration before absolute risk estimation. HHD yielded the highest AUC: XGBoost on the balanced dataset achieved 0.758 (95% CI, 0.735 to 0.777), classified as high utility. The Super Learner ensemble was competitive for HHD on the unbalanced dataset (0.754; 95% CI, 0.731 to 0.776). AF/AFL followed at 0.732 (95% CI, 0.715 to 0.750; XGBoost, balanced), representing moderate utility. HF achieved 0.708 (95% CI, 0.701 to 0.716; XGBoost, balanced), with the narrow confidence interval driven by the large test sample (28,380 records from 21,149 patients). AMI achieved 0.706 (95% CI, 0.691 to 0.721; XGBoost, unbalanced). XGBoost was the best-performing base algorithm across all four categories. ROC curves are shown in **Figure 2**; the cross-category algorithm comparison appears in **Figure 3**.

AUPRC values further characterized discrimination in the context of varying event rates: HF 0.418 (event rate 22.5%), AF/AFL 0.339 (15.3%), HHD 0.338 (12.4%), and AMI 0.275 (13.0%).

The relatively higher AUPRC for HF reflects the higher base rate, while AMI’s lower AUPRC despite comparable AUC suggests that positive prediction remains challenging for the least prevalent readmission category.

### Effect of SMOTE Balancing

Balanced XGBoost achieved the best AUC for HHD (0.758) and HF (0.708), while unbalanced XGBoost was best for AMI (0.706). AF/AFL showed virtually no difference between balanced and unbalanced XGBoost (0.732 vs 0.731). Across categories, SMOTE generally improved sensitivity at the expense of specificity, a tradeoff that was most beneficial in categories with lower event rates (HHD 12.4%, AMI 13.0%; **Supplementary Table S9**).

### Model Calibration

Calibration was assessed using Brier scores, calibration slopes, and E/O ratios (**Figure 4**). The AMI XGBoost model demonstrated excellent relative calibration (slope = 0.993), indicating well-preserved rank ordering of predicted probabilities. However, the expected-to-observed (E/O) ratio of 3.47 indicates substantial overestimation of absolute risk, meaning the model predicted 3.47 times more readmission events than were observed. While the model reliably identifies higher-risk patients relative to lower-risk patients, absolute probability estimates for AMI require recalibration (e.g., Platt scaling or isotonic regression) before clinical deployment. HF models showed excellent calibration (balanced slope 1.014, unbalanced slope 1.025), with Brier scores of 0.158 (balanced) and 0.215 (unbalanced). AF/AFL exhibited slight underconfidence (slopes 1.037–1.065). HHD showed the greatest miscalibration (slopes 1.479–1.646), indicating systematic overconfidence at the extremes of predicted risk; Platt recalibration is recommended before deployment for this subtype. Raw Brier scores are reported in **Table 2**; for all categories, rank-ordering of risk was preserved even when probability estimates required rescaling.

### Pairwise Algorithm Comparisons

DeLong tests for correlated ROC curves compared all algorithm pairs within the best-performing dataset configuration per CVD category (**Supplementary Table S3**). XGBoost significantly outperformed Random Forest and Elastic Net across most conditions. Within each condition, XGBoost and LightGBM were generally statistically indistinguishable, though XGBoost held a consistent numerical edge. The Super Learner was competitive with or slightly below the best base learner across all categories, with the closest match in HHD.

### Patient-Grouped Validation

GEE models with exchangeable working correlation and sandwich standard errors were fitted to the multi-admission subset of each category’s test partition (**Supplementary Table S4**). The GEE coefficient for predicted risk was significant in HF (α=0.120, Wald P<0.001), AMI (α=0.041, Wald P<0.001), and HHD (α=0.141, Wald P=0.017), confirming that model-predicted risk remained a significant readmission predictor after accounting for within-patient correlation. AF/AFL showed a non-significant correlation (α=0.170, P=0.229), likely due to the smaller multi-admission sample (1,067 records from 461 patients).

Clustered bootstrap resampling (1,000 iterations resampling patients with replacement) produced confidence intervals comparable to standard record-level bootstrap intervals (**Supplementary Table S5**). The mean CI width ratio across categories was 1.13; patient clustering widened intervals by approximately 13% on average. No model’s point estimate fell outside its clustered interval, and all utility classifications remained unchanged. Taken together, the patient-grouped splitting strategy adequately controlled within-patient correlation.

The clustered bootstrap confidence interval for HHD (0.733–0.784) extends below the 0.75 threshold defining “high utility” in our classification schema, despite the point estimate (0.758) exceeding this threshold. The 21.2% CI widening from patient-level clustering for HHD was the largest among all four categories, reflecting greater within-patient correlation in this subgroup. The “high utility” classification for HHD should therefore be interpreted with this uncertainty in mind, particularly when considering prospective deployment.

### Performance by Admission Order

To assess whether prior hospitalization history influenced discrimination, best-model AUC was stratified by admission order within each test partition (**Supplementary Table S6**). Discrimination varied by admission history, with greater heterogeneity among repeat admissions in categories with lower event rates. The Super Learner provided the most stable performance across admission strata.

LACE Index Sensitivity Analysis. To assess the impact of construct overlap between the LACE Index composite score and its constituent components (LACE-L, LACE-A, LACE-C, LACE-E), we conducted a sensitivity analysis comparing three feature configurations using XGBoost: (1) the full consensus feature set including both the composite and components, (2) LACE composite index only (individual components removed), and (3) LACE components only (composite removed). AUC differences were negligible across configurations (**Supplementary Table S11).** For AMI, HF, and AF/AFL, all three configurations produced identical AUCs to the third decimal place (all DeLong p > 0.20). For HHD, the composite-only configuration yielded marginally higher AUC (0.755) than both the full set (0.747, DeLong p = 0.053) and components-only (0.747, p = 0.015), suggesting the LACE Index composite alone is sufficient and may even reduce noise from redundant component features. These results support using the composite LACE Index alone for parsimony and clinical interpretability, and confirm that construct overlap did not artificially inflate model discrimination.

### Feature Importance

Aggregated importance across all 32 base models produced a stable predictor ranking (**Figure 5**, **Supplementary Table S7**). Validated clinical indices dominated: the LACE Index ranked among the top predictors in every category, in line with its strong separation between readmitted and non-readmitted patients (SMD=0.604). The Charlson Comorbidity Index, van Walraven Elixhauser Index, and Elixhauser Comorbidity Count also ranked highly, a pattern that follows the shift from ad-hoc composites to published indices with established reference ranges.

Category-specific patterns supported the case for condition-specific modeling (**Figure 5**). In HF, the LACE Index and renal failure dominated, with admission order contributing strongly. In AMI, total diagnoses and the LACE Index led. In AF/AFL, the importance distribution was broader, with insurance type contributing more prominently. In HHD, the LACE Index, insurance type, and renal failure led, with obesity and depression contributing more than in other categories. Renal failure showed consistent importance across all categories (SMD=0.325 for readmission vs non-readmission), in keeping with the cardiorenal syndrome’s known role in CVD readmission.

Elastic Net coefficient vectors (**Supplementary Table 7**) offered a view of predictor direction and regularization-driven sparsity. The LACE Index and renal failure carried the largest positive coefficients across conditions, while obesity consistently received negative coefficients, suggesting a protective association after adjustment for comorbidity burden.

SHAP analysis replicated the global importance rankings at the individual prediction level (**Figure 6**). The LACE Index, admission order, renal failure, and insurance type produced the largest absolute SHAP values across predictions. Waterfall decompositions of representative cases (**Supplementary Figure S4**) illustrated how these top features combined to produce individual risk estimates. Dependence plots (**Supplementary Figure S3**) revealed non-linear relationships, including threshold effects for the LACE Index and differential effects of insurance type across predicted risk levels.

## Discussion

This study is, to our knowledge, the largest ML-based cardiovascular readmission prediction analysis in a predominantly Black cohort and the first to compare five algorithms plus a Super Learner ensemble across four CVD subtypes using standardized validated features in a racially concentrated population.

### Condition-Specific Discrimination

All four CVD categories reached moderate or high predictive utility, exceeding most prior administrative data-based benchmarks. The HF AUC of 0.708 exceeded both Sharma and colleagues’ benchmark of 0.65 using XGBoost on Canadian administrative data^27^ and Mortazavi and colleagues’ Random Forest AUC of 0.628 in the Tele-HF clinical trial cohort,^39^ suggesting that validated indices and consensus feature selection narrow the discrimination gap previously attributed to administrative data limitations. Fine and colleagues achieved 0.74 for a composite HF endpoint using CatBoost with deep feature synthesis, though their outcome included ED revisits and mortality.^28^ Awan and colleagues similarly reported a best AUC of 0.62 for HF readmission or death using administrative data with explicit attention to class imbalance.^18^

The HHD model reached high utility (AUC 0.758). Because most studies subsume HHD within broader HF or hypertension categories,^1,2^ direct comparison with published benchmarks is limited. The narrow AUC range across algorithms for HHD likely reflects a more uniform pathophysiology, making this subtype a practical candidate for early deployment. AMI models achieved moderate utility (0.706) with the best calibration in the study (slope 0.993), within the range of published administrative data AMI models (AUC 0.65–0.75 without clinical variables).^6,7^ Pandey and colleagues found that racial differences in AMI readmission were attributable to patient-level rather than hospital-level factors,^12^ which aligns with the strong insurance and comorbidity contributions in our AMI models. AF/AFL reached moderate utility (0.732), likely because validated indices better captured the comorbidity burden driving arrhythmia-related readmissions.

### Algorithmic Hierarchy

XGBoost outperformed all alternatives across four categories, extending results from mixed-race populations^5,17,19^ to a predominantly Black cohort with identical features. The Super Learner ensemble matched the best base learner most closely for HHD (unbalanced AUC 0.754 vs XGBoost 0.753), where LightGBM and Ranger received the largest meta-learner weights (**Supplementary Figure S2**), suggesting that ensemble gains require base learners with complementary decision boundaries. Balanced datasets produced the best AUC in two of four categories (HHD, HF), with unbalanced best for AMI and no meaningful difference for AF/AFL.

### Insurance, Social Determinants, and Health Equity

The LACE Index was the strongest discriminator between readmitted and non-readmitted patients (SMD=0.604) and, unlike ad-hoc composite scores, has established reference ranges and external validity across settings.^32^ Insurance type ranked among the top predictors (SMD=0.342). Bahiru and colleagues similarly showed that hospitals serving higher proportions of dual-eligible patients had significantly higher 30-day readmission rates.^30^ The low readmission representation among self-pay and uninsured discharges (5.3% vs 10.7%) more likely reflects access barriers to rehospitalization than better outcomes,^11,14,22^ meaning ML models trained on these data may underestimate true risk for uninsured patients. Chaiyachati and colleagues showed that racial disparities in readmission widened within safety-net hospitals after HRRP implementation, especially for conditions not targeted by HRRP,^31^ adding weight to the case that HRRP risk adjustment models should incorporate social determinant measures. Condition-specific feature importance patterns, such as the prominent role of obesity and depression in HHD, would have been missed in pooled modeling.

Predictive contribution is distinct from clinical actionability. The LACE Index, renal failure, and obesity improve risk stratification but are not directly modifiable in the acute window. Insurance type is partly modifiable through policy yet also proxies for age, disability, and post-discharge access. Care process features (admission order, length of stay, hospital characteristics) identify where targeted transition-of-care interventions may reduce readmission probability. Model outputs should be interpreted accordingly: non-modifiable features refine risk classification, while care process features indicate intervention opportunities.

### Feature Importance Concordance

Algorithm-native, aggregated, and SHAP-based importance rankings all placed the LACE Index, renal failure, and insurance type among the top predictors, paralleling the emphasis on appropriate model and metric selection for administrative HF readmission data reported by Awan and colleagues.^18^ The LACE Index, Charlson, and van Walraven Elixhauser indices carry published reference ranges and established predictive validity, making them more reproducible and interpretable than ad-hoc composites.

However, the LACE Index requires careful interpretation as a predictor because its components (length of stay, acuity of admission, comorbidity, and emergency department visits) overlap with readmission risk factors, creating potential construct overlap. While this does not invalidate its predictive utility, it means that LACE-based risk stratification identifies high-utilization patients through a partially tautological construct. SHAP dependence plots (**Supplementary Figure S3**) revealed non-linear LACE–readmission relationships, including diminishing marginal effects at the highest scores.

Our sensitivity analysis (**Supplementary Table S11**) confirmed that the simultaneous inclusion of the LACE Index and its constituent components did not inflate model discrimination: AUC was unchanged across all three feature configurations for AMI, HF, and AF/AFL (all DeLong p > 0.20), and the composite-only configuration actually performed marginally better for HHD (DeLong p = 0.015 vs. components-only). The composite LACE Index alone is therefore sufficient for prediction, and its high feature importance ranking reflects genuine predictive value rather than redundant information from overlapping components.

### Practical Implications

All four CVD models demonstrated moderate to high utility for risk stratification, supporting their integration into clinical workflows. The HHD model (AUC 0.758, high utility) is the most immediate candidate for pilot clinical decision support in safety-net settings. HF and AF/AFL models, now at moderate utility, can support tiered risk stratification within the planned Readmission Analytics and Interactive Platform (RAIP) dashboard. Low PPVs across all models, as seen in comparable studies,^27,28^ indicate that integration within a tiered risk stratification system rather than standalone screening remains appropriate. The Super Learner’s competitive performance for HHD provides an ensemble option for deployment contexts where model stability is valued over marginal discrimination gains.

The HHD model shows high discrimination but suboptimal calibration (slopes 1.479-1.646), suggesting that predicted probabilities at extremes are overstated. Thus, it is suitable for ranking patients by risk but requires recalibration, via Platt scaling or isotonic regression, before absolute risk communication.

Key strengths include a large, predominantly Black cohort (157,791 discharges, 96.6% Black), a fully crossed 40-model design enabling unbiased algorithm comparison, data-driven consensus feature selection, and validated clinical indices for reproducibility.^14,16,20^ Patient-grouped splitting with GEE post-hoc validation and clustered bootstrap resampling ensured adequate control for within-patient correlation. Identifying HHD as a standalone prediction target with the highest discrimination offers an actionable result for a condition routinely subsumed within broader categories.

Several limitations warrant discussion. The 96.6% Black cohort limits generalizability to diverse populations and requires external validation.^16,20^

Lack of laboratory values, vital signs, ejection fraction, medications, and four key administrative features (APR-DRG Severity, Risk of Mortality, Admission Type, Discharge Disposition) may have constrained model performance. ICD-based classification cannot distinguish HFpEF from HFrEF. Low readmission among self-pay/uninsured patients introduces potential bias, as observed outcomes may underestimate true clinical risk. SMOTE-generated synthetic observations, particularly for binary comorbidity flags, may not fully reflect the true feature space, and alternative resampling strategies (e.g., SMOTE-NC or conditional generative models) may better preserve data fidelity. Model selection from multiple candidates likely overestimates performance (optimistic bias), emphasizing the need for independent external validation (Supplementary Table S3). The LACE Index, as both a predictor and a construct overlapping with readmission determinants, introduces potential circularity that warrants evaluation in external validation.

The ICD-9 to ICD-10 transition may have introduced systematic differences in comorbidity ascertainment over the study period, affecting feature distributions despite adjustment for admission year (Supplementary Table S10). Future studies should consider multi-site validation and stratification by coding era to mitigate these temporal effects.

Finally, another limitation is the absence of independent external validation. Although models were evaluated using a patient-grouped 70/30 train–test split with additional robustness checks including clustered bootstrap resampling and GEE analyses to account for within-patient correlation, these procedures represent internal validation within a single statewide dataset.

Consequently, model performance estimates may remain optimistic relative to deployment in new clinical environments. Administrative coding practices, case mix, health system structure, and social determinants vary across regions and institutions, and these factors may influence both predictor distributions and readmission risk relationships. Therefore, the present models should be interpreted as development-stage prediction tools rather than finalized clinical decision support systems. Independent validation in geographically distinct populations and healthcare systems will be necessary to assess generalizability and confirm model calibration and discrimination

## Conclusions

Condition-specific ML models achieved moderate-to-high discrimination for 30-day readmission across four CVD subtypes in a large, predominantly Black cohort. Model performance was driven more by CVD subtype than algorithm choice, with XGBoost performing consistently and the Super Learner competitive for HHD. Key predictors, including the LACE Index, renal failure, and insurance status, highlight the combined influence of clinical and structural factors. These results support deploying subtype-specific, equity-informed readmission models using validated clinical indices rather than ad-hoc composites.

External validation in diverse populations, incorporation of area-level social determinants, and prospective evaluation of model-guided interventions, particularly for HHD and HF, are needed to confirm generalizability and assess impact on readmission disparities.

## Acknowledgments

This research was supported by American Heart Association Second Century Implementation Science Award #23SCISA1145640. The opinions expressed in this article are solely those of the authors and do not necessarily represent those of the American Heart Association. This project was initiated at the EVMS-Sentara Healthcare Analytics and Delivery Science Institute (HADSI), Eastern Virginia Medical School, and continued at the Macon & Joan Brock Virginia Health Sciences at Old Dominion University following the institutional integration of EVMS and Old Dominion University. The authors acknowledge collaborative research support provided through a subaward to the University of Illinois College of Medicine, Peoria, IL. Data access was facilitated by the M. Foscue Brock Institute for Community and Global Health and the Research and Infrastructure Service Enterprise (RISE) at the Macon & Joan Brock Virginia Health Sciences, Old Dominion University. Virginia Health Information (VHI) has provided non-confidential patient-level information used in this study, which it has compiled in accordance with Virginia law but which it has no authority to independently verify. By using this data, the authors agree to assume all risks that may be associated with or arise from the use of inaccurate data. VHI cannot and does not represent that the use of VHI’s data was appropriate for this study or endorse or support any conclusions or inferences that may be drawn from the use of VHI’s data. This study was approved by the Eastern Virginia Medical School Institutional Review Board (IRB #23-09-NH-0248) with a waiver of informed consent for de-identified data analysis.

## Sources of Funding

This research was funded by the American Heart Association Second Century Implementation Science Award (Award #23SCISA1145640; PI: Ismail El Moudden). The funder had no role in the design of the study; in the collection, analysis, or interpretation of data; in the writing of the manuscript; or in the decision to submit the article for publication.

## Disclosures

Dr. El Moudden reports research funding from the American Heart Association (Award #23SCISA1145640) during the conduct of this study; no other relationships or activities that could appear to have influenced the submitted work. Dr. Dodani reports the following relationships: Vice-Chair, American Heart Association Health Equity Research Network (HERN) Oversight Advisory Committee; subaward through the University of Illinois at Chicago for collaborative research support on this project (Award #23SCISA1145640). She holds appointments at the University of Illinois College of Medicine–Peoria (Founding Director, Center 4 Health Research; Professor of Clinical Medicine, Department of Medicine) and at the Macon & Joan Brock Virginia Health Sciences, Eastern Virginia Medical School at Old Dominion University (Professor of Medicine, Community Non-Tenure Track). No other relationships or activities that could appear to have influenced the submitted work. Mr. Bittner reports no disclosures.

## Non-standard Abbreviations and Acronyms

AF/AFL: atrial fibrillation/flutter
AMI: acute myocardial infarction
AUC: area under the receiver operating characteristic curve
AUPRC: area under the precision-recall curve
CMS: Centers for Medicare & Medicaid Services
GEE: generalized estimating equations
HHD: hypertensive heart disease
HRRP: Hospital Readmissions Reduction Program
LACE: Length of stay, Acuity, Comorbidities, Emergency department visits
LightGBM: Light Gradient Boosting Machine
ML: machine learning
mRMR: minimum redundancy maximum relevance
SDOH: social determinants of health
SHAP: SHapley Additive exPlanations
SMOTE: Synthetic Minority Oversampling Technique
TRIPOD: Transparent Reporting of a multivariable prediction model for Individual Prognosis Or Diagnosis
VHI: Virginia Health Information XGBoost Extreme Gradient Boosting

## Notes

### Clinical Trial

Not applicable. This is a retrospective cohort study using de-identified administrative discharge data and does not meet the definition of a clinical trial requiring prospective registration.

### Author Declarations

Eastern Virginia Medical School Institutional Review Board, IRB #23-09-NH-0248. Approval granted with waiver of informed consent for de-identified data analysis.

